# Epidemiological and microbiological investigation of the large increase of vibriosis in northern Europe in 2018

**DOI:** 10.1101/2021.11.19.21266449

**Authors:** Ettore Amato, Maximilian Riess, Daniel Thomas-Lopez, Marius Linkevicius, Tarja Pitkänen, Tomasz Wołkowicz, Jelena Rjabinina, Cecilia Jernberg, Marika Hjertqvist, Emily MacDonald, Jeevan Karloss Antony-Samy, Karsten Dalsgaard Bjerre, Saara Salmenlinna, Kurt Fuursted, Anette Hansen, Umaer Naseer

**Affiliations:** Department of Infection Control and Preparedness, Norwegian Institute of Public Health, Oslo, Norway; Department of Microbiology, Public Health Agency of Sweden, Department of Microbiology, Stockholm, Sweden; Department of Bacteria, Parasites & Fungi, Division of Infectious Disease Preparedness, Statens Serum Institut, Copenhagen, Denmark; Finnish Institute for Health and Welfare, Department of Health Security, Helsinki, Finland; European Programme for Public Health Microbiology Training (EUPHEM), European Centre for Disease Prevention and Control (ECDC), Stockholm, Sweden; Finnish Institute for Health and Welfare, Department of Health Security, Kuopio, Finland; University of Helsinki, Helsinki, Finland; National Institute of Public Health, Warsaw, Poland; Health Board, Department of CD Surveillance and Control, Tallinn, Estonia; Public Health Agency of Sweden, Department of Communicable Disease Control and Health Protection, Stockholm, Sweden; Data Integration and Analysis, Division of Infectious Disease Preparedness, Statens Serum Institut, Copenhagen, Denmark

**Keywords:** vibriosis, seawater, emerging pathogens, heatwaves, global warming

## Abstract

Northern European countries and countries bordering the Baltic Sea have witnessed an increase of vibriosis cases during recent heatwaves. Here, we described the epidemiology of vibriosis cases and the genetic diversity of *Vibrio* isolates from Norway, Sweden, Denmark, Finland, Poland, Estonia, and Latvia in 2018, a year with an exceptionally warm summer. We conducted a retrospective study and analysed demographics, geographic distribution, seasonality, causative species, and severity of non-travel related vibriosis cases in 2018. Data sources included surveillance systems, national laboratory notification databases and/or nationwide surveys to public health microbiology laboratories. Moreover, we performed whole genome sequencing and multilocus sequence typing of available isolates from 2014-2018 to map their genetic diversity.

In 2018, we identified 445 non-travel related vibriosis cases in the study countries, which was considerably higher than the median of 126 cases between 2014-2017 (range: 87-272). The main reported mode of transmission was exposure to seawater. We observed a species-specific geographic disparity of vibriosis cases across the Nordic-Baltic region. Developing severe vibriosis was associated with infections of *Vibrio vulnificus* (adjOR: 17.2; 95% CI: 3.3-90.5) or *Vibrio parahaemolyticus* (adjOR: 2.1; 95% CI: 1.0-4.5), being ≥65 years of age (65-79 years, adjOR: 3.9; 95% CI: 1.7-8.7; 80+ years, adjOR: 15.5; 95% CI: 4.4-54.3) or acquiring infections during summer (adjOR: 5.1; 95% CI: 2.4-10.9). Although phylogenetic analysis revealed diversity between *Vibrio* isolates, two *V. vulnificus* clusters (<10 SNPs) were identified. A share sentinel surveillance system for vibriosis during summer months might be highly valuable to monitor this emerging public health issue.

## Introduction

The habitat of *Vibrio* bacteria is fresh and brackish water with moderate salinity. Non-toxigenic *Vibrio cholerae*, as well as several human pathogenic non-cholera *Vibrio* species, including *Vibrio alginolyticus, Vibrio parahaemolyticus* and *Vibrio vulnificus*, cause vibriosis after seawater exposure or consumption of contaminated seafood [1]. Clinical manifestations range from mild gastroenteritis and otitis to wound infections that may lead to severe necrotizing fasciitis and septicaemia with a potentially fatal outcome [2-5]. The Baltic Sea region is one of the areas where increasing numbers of cases related to *Vibrio* species causing vibriosis (VCV) have been reported in the last decades [6]. Several studies have shown how the occurrence of heatwaves, that lead to an increase of sea surface temperature, are linked with an increase of the number of reported vibriosis cases [4, 7-12]. For instance, 2006, 2010, and particularly 2014 (the warmest year in historical records to that date), were the years with a significantly warm summer in the Baltic Sea region, and were also the years with the largest number of vibriosis cases reported [6, 11]. However, there is a notable gap in surveillance data of vibriosis [1, 6], since it is not a notifiable disease in the majority of European countries. Therefore, the aim of this multi-country study was to describe the epidemiology of vibriosis cases in countries bordering the North and Baltic Seas area during the most recent exceptionally warm year of 2018 [13, 14], in order to investigate the extension of these infections in the study countries, map their genetic diversity, understand the predictors for developing severe vibriosis, and to propose recommendations for public health measures.

## Methods

### Study design and case definition

We conducted a retrospective cross-sectional study to analyse the epidemiology of VCV infections reported in 2018 in Norway, Denmark, Sweden, Finland, Poland, Estonia, and Latvia, further referred to as the study countries. Available data of vibriosis cases since the last warmest summer (2014) were used to contextualize the number of VCV infections of 2018.

A case of vibriosis was defined as a laboratory confirmed VCV infection from the study countries; those related to travel were excluded. For few cases (n=18) more than one *Vibrio* species were recorded concurrently in the same patient. In such cases only the species and sample type related to a more severe infection was included.

### Data source and collection

Each country used different data sources including compulsory comprehensive passive surveillance systems for vibriosis (Sweden, Finland, Poland, and Estonia), national laboratory notification databases (Denmark and Latvia) or nationwide surveys to public health microbiology laboratories (Norway) (Table S1).

The reporting criteria varied between countries that had a surveillance system in place in 2018. In Sweden, a confirmed case was defined as an isolation of *Vibrio* spp. other than toxigenic *V. cholerae* O1 or O139. In Finland, a case was defined as (i) *V. cholerae* including non-O1, non-O139 from faecal sample, culture or polymerase chain reaction (PCR) (or other nucleic acid detection); (ii) *V. parahaemolyticus* from faecal sample, culture or PCR (or other nucleic acid detection) and (iii) any *Vibrio* spp. from blood sample or cerebrospinal fluid, culture or PCR (or other nucleic acid detection). In Poland, a case was defined according to the International Classification of Diseases 10^th^ revision, diagnosis A05.3 for *V. parahaemolyticus*. In Estonia, a vibriosis case was considered as any case meeting the clinical criteria (otitis, wound infection, gastroenteritis, septicaemia) and laboratory criteria (detection of *Vibrio* spp., *V. cholerae* non-O1, non-O139 in a clinical specimen detected by any method). Meanwhile, the criteria for *Vibrio* spp. infections reported from national laboratory notification databases (Denmark) and laboratory nationwide surveys (Norway) were based on detection of *Vibrio* spp. other than toxigenic *V. cholerae* O1 or O139. No surveillance system for vibriosis was in place in Latvia.

We compiled the vibriosis cases from all study countries into a harmonized dataset that included: patients’ sex, age group, year and month of infection, country, European NUTS3 (nomenclature of territorial units for statistics 3) region [15], identified VCV, type of sample and, if known, source of exposure and travel status at the probable time of infection. The severity of an infection was inferred from the sample type: blood/serum (n=60) and wound swabs (n=144) were considered as a proxy of severe infections, while skin swabs (n=28), ear secretion (n=176), faeces (n=19), urine (n=2), nasal swab (n=1) and other unspecified (n=15) sample types were considered linked to non-severe infections. Seasons were defined according to the northern hemisphere seasons (spring: March – May; summer: June – August; autumn: September – November; winter: December – February). Population data as per 31 December 2018 were publicly available from national statistics authorities.

### Epidemiological investigation and statistical analysis

We described the epidemiology of vibriosis cases reported in 2018 in the study countries per country and as total counts. Data presented included the sex-ratio, notification rate per 100,000 inhabitants, median age, distribution of cases across age groups, season and identified VCV (Table S2). Case numbers were presented by country and by region (NUTS3) while seasonality was further depicted by month of infection. Severity of infection was described by age group, and month of infection. Association of sex, age group, season, and VCV with developing severe vibriosis was further analysed by estimation of crude odds ratios (OR) and 95% confidence intervals (CI) by univariate logistic regression analysis. Adjusted OR (adjOR) with 95% CI were estimated in a multivariable analysis. Binary outcome was severe/non-severe vibriosis.

Data analysis was performed using Stata version 15.0 (2017. Stata Statistical Software: Release 15. College Station, TX: StataCorp LP. USA). Categorical variables were described as proportions with 95% CI and were compared using chi-squared test. Continuous variables were described using mean and standard deviation or median and range, and were compared using t-test or non-parametric Wilcoxon rank-sum test. Trends were assessed using a nonparametric test across ordered groups. Observations with missing values for variables under comparison were excluded from the respective analysis. We used an alpha level of 0.05 for all statistical tests. Stata outputs of p-values p<0.000 are reported as p<0.001.

### Sampling of VCV isolates, MLST and WGS analyses

We collected available clinical VCV isolates in 2018 from the national public health institutes or regional laboratories, and complemented them with available clinical (2014-2017) and environmental (2018) isolates (Table S3). DNA was extracted and sequenced using standard operating procedures and Illumina sequencers. WGS raw files are available at the European Nucleotide Archive (https://www.ebi.ac.uk/ena) under study project accession number PRJEB43461. Accession numbers of all sequenced isolates are listed in Table S4.

Raw reads from each country were analysed together using a common pipeline for species identification, MLST, and phylogenetic analyses. We used BBmap (version 38.69) to clean the raw reads and FastQC (version 0.11.8) to generate quality reports of samples. Additionally, we used Kraken2 (version 2.0.8_beta) to confirm the species and Shovill (1.0.9) to assemble (using SPAdes version 3.13.1) the genomes.

We searched the PubMLST database (https://pubmlst.org/) using Ariba (2.14.4). Assignment of sequence type (ST) was performed for isolates of non-toxigenic *V. cholerae, V. parahaemolyticus, V. vulnificus* and *V. alginolyticus* according to their respective MLST schemes (Figure 4 and 5, S1-S2).

We used Parsnp (v1.2) and a neighbour-joining algorithm to build the phylogenetic trees, and Snp-dists (0.7.0) to calculate the single nucleotide polymorphism (SNP) distance between isolates. A cluster was defined as two or more isolates within 30 SNPs difference. An in-house pipeline was used for sequence mapping, generation of consensus sequences, alignment calculation, and SNP filtering (exclusion distance = 300). We used R package ggtree [16] to visualise the phylogenetic trees generated by the in-house pipeline (https://github.com/folkehelseinstituttet/Vibrio-Project).

## Results

### Descriptive epidemiology of vibriosis cases

In 2018, 445 non-travel related cases of vibriosis were reported in the study countries, which was the highest single year case number compared to the four previous years (n=610) (Figure 1A, Table 1 and S2).

**Table 1.** Summary of epidemiological parameters of vibriosis cases per species in the study countries, 2018.

|  |  | Total vibriosis (N=445) |  | Vibrio alginolyticus (N=152) |  | Non-toxicogenic Vibrio cholerae (N=100) |  | Vibrio parahaemolyticus (N=89) |  | Vibrio vulnificus (N=45) |  | Non-subtyped Vibrio spp. (N=59) |  |
| --- | --- | --- | --- | --- | --- | --- | --- | --- | --- | --- | --- | --- | --- |
|  |  | n | % | n | % | n | % | n | % | n | % | n | % |
| Sex | Female | 168 | 37.8 | 68 | 45.0 | 26 | 26.0 | 38 | 43.0 | 14 | 31.0 | 22 | 37.3 |
|  | Male | 277 | 62.2 | 84 | 55.0 | 74 | 74.0 | 51 | 57.0 | 31 | 69.0 | 37 | 62.7 |
| Age group | 0-4 | 9 | 2 | 2 | 1.3 | 4 | 4.0 | 0 | 0.0 | 1 | 2.2 | 2 | 3.4 |
|  | 5-14 | 91 | 20.4 | 47 | 30.9 | 23 | 23.0 | 7 | 7.9 | 0 | 0.0 | 14 | 23.7 |
|  | 15-24 | 47 | 10.6 | 24 | 15.8 | 10 | 10.0 | 4 | 4.5 | 0 | 0.0 | 9 | 15.3 |
|  | 25-44 | 54 | 12.1 | 26 | 17.1 | 12 | 12.0 | 7 | 7.9 | 1 | 2.2 | 8 | 13.6 |
|  | 45-64 | 83 | 18.7 | 24 | 15.8 | 24 | 24.0 | 20 | 22.5 | 6 | 13.3 | 9 | 15.3 |
|  | 65-79 | 109 | 24.5 | 24 | 15.8 | 19 | 19.0 | 36 | 40.5 | 21 | 46.7 | 9 | 15.3 |
|  | >80 | 52 | 11.7 | 5 | 3.3 | 8 | 8.0 | 15 | 16.9 | 16 | 35.6 | 8 | 13.6 |
| Season | Summer | 326 | 73.3 | 97 | 63.8 | 74 | 74.0 | 78 | 87.6 | 44 | 97.8 | 33 | 55.9 |
|  | Autumn | 96 | 21.6 | 45 | 29.6 | 22 | 22.0 | 6 | 6.7 | 1 | 2.2 | 22 | 37.3 |
|  | Winter | 13 | 2.9 | 6 | 4.0 | 2 | 2.0 | 3 | 3.4 | 0 | 0.0 | 2 | 3.4 |
|  | Spring | 10 | 2.2 | 4 | 2.6 | 2 | 2.0 | 2 | 2.3 | 0 | 0.0 | 2 | 3.4 |
| Country | Norway | 92 | 20.7 | 63 | 41.5 | 2 | 2.0 | 12 | 13.5 | 9 | 20.0 | 6 | 10.2 |
|  | Denmark | 170 | 38.2 | 70 | 46.1 | 3 | 3.0 | 55 | 61.8 | 16 | 35.6 | 26 | 44.1 |
|  | Sweden | 147 | 33 | 19 | 12.5 | 64 | 64.0 | 19 | 21.4 | 19 | 42.2 | 26 | 44.1 |
|  | Finland | 30 | 6.7 | 0 | 0.0 | 26 | 26.0 | 3 | 3.4 | 1 | 2.2 | 0 | 0.0 |
|  | Poland, Estonia | 6 | 1.3 | 0 | 0.0 | 5 | 5.0 | 0 | 0.0 | 0 | 0.0 | 1 | 1.7 |
| Sample type | Blood | 60 | 13.5 | 3 | 2.0 | 20 | 20.0 | 4 | 4.5 | 31 | 68.9 | 2 | 3.4 |
|  | Faeces | 19 | 4.3 | 2 | 1.3 | 11 | 11.0 | 3 | 3.4 | 0 | 0.0 | 3 | 5.1 |
|  | Ear-related | 176 | 39.6 | 91 | 59.9 | 43 | 43.0 | 14 | 15.7 | 1 | 2.2 | 27 | 45.8 |
|  | Wound-related | 144 | 32.4 | 45 | 29.6 | 13 | 13.0 | 54 | 60.7 | 12 | 26.7 | 20 | 33.9 |
|  | Other | 46 | 10.3 | 11 | 7.2 | 13 | 13.0 | 14 | 15.7 | 1 | 2.2 | 7 | 11.9 |
| Exposure | Food/water | 6 | 1.3 | 2 | 1.3 | 3 | 3.0 | 1 | 1.1 | 0 | 0.0 | 0 | 0.0 |
|  | Bathing/seawater | 109 | 24.5 | 17 | 11.2 | 38 | 38.0 | 12 | 13.5 | 25 | 55.6 | 17 | 28.8 |
|  | Other | 1 | 0.2 | 1 | 0.7 | 0 | 0.0 | 0 | 0.0 | 0 | 0.0 | 0 | 0.0 |
|  | Unknown | 329 | 73.9 | 132 | 86.8 | 59 | 59.0 | 76 | 85.4 | 20 | 44.4 | 42 | 71.2 |
| Severe infection | Yes | 204 | 45.8 | 48 | 31.6 | 33 | 33.0 | 58 | 65.2 | 43 | 95.6 | 22 | 37.3 |
|  | No | 241 | 54.2 | 104 | 68.4 | 67 | 67.0 | 31 | 34.8 | 2 | 4.4 | 37 | 62.7 |

Note: No cases were reported for Latvia.

**Figure 1.**
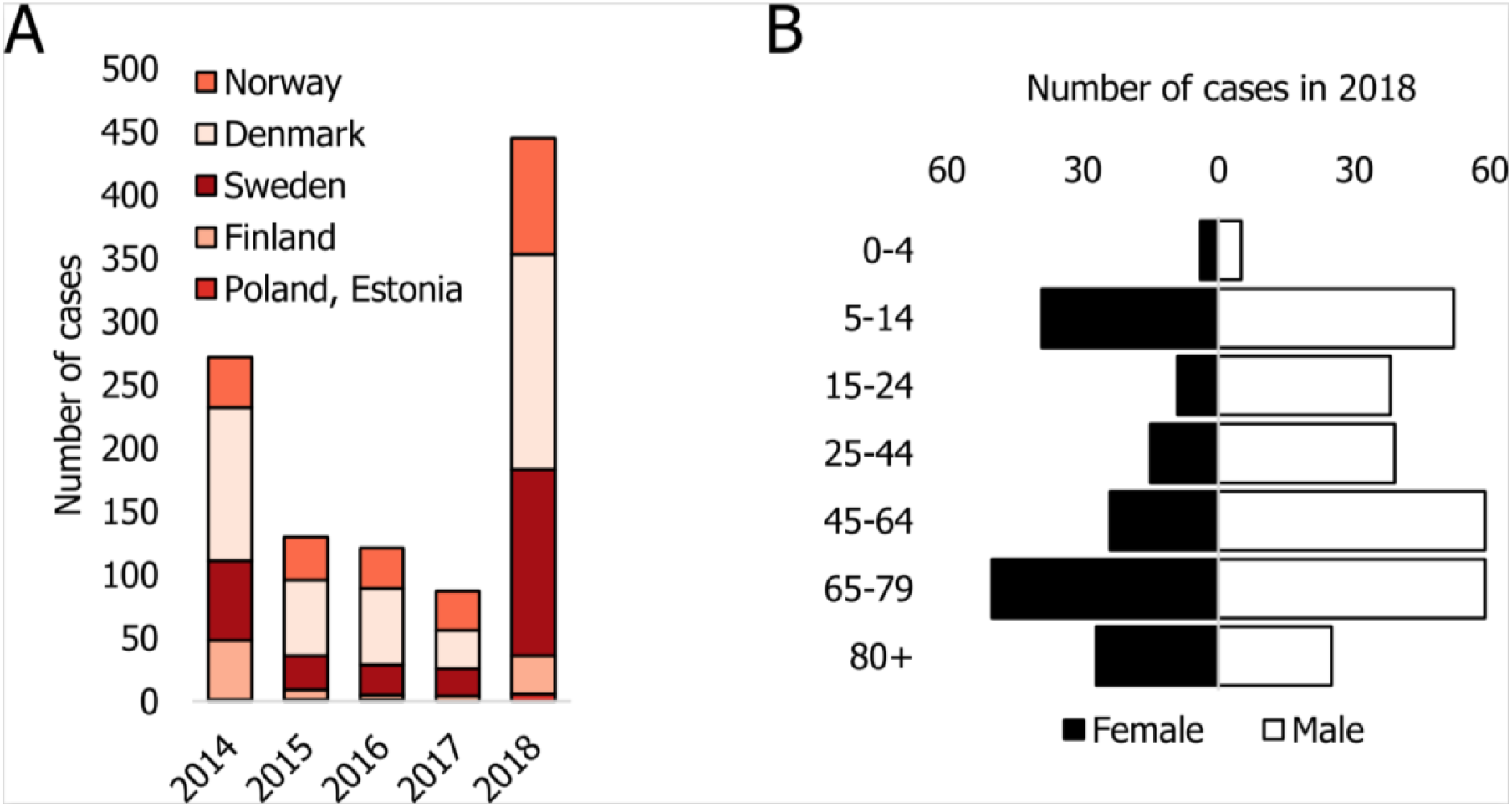
Occurrence of vibriosis cases in study countries during 2014-2018 (A) and distribution of cases by age and sex (B) in 2018.

The vibriosis notification rates ranged between 0.5 in Finland and 2.9 per 100,000 inhabitants in Denmark. Due to limited number of cases (n=6), the notification rate was not calculated for Poland and Estonia. Latvia reported no cases (Table S2). The majority of the cases were male (n=277, 62.2%) (Table 1) and the highest number of cases was reported in the age group 65-79 (n=109, 24.5%) followed by age groups 5-14 (n=91, 20.4%) and 45-64 years old (n=83, 18.7%) (Figure 1B, Table 1).

Most of the infections were caused by *V. alginolyticus* (n=152, 34.2%), followed by non-toxigenic *V. cholerae* (n=100, 22.5%), *V. parahaemolyticus* (n=89, 20%), *V. vulnificus* (n=45, 10.1%), and non-subtyped *Vibrio* spp. (n=59, 13.2%). The most common type of infections reported were ear infections (n=176; 39.6%), followed by wound infections (n=144; 32.4%) (Table 1).

We observed a difference in the proportions of species affecting each age group. The proportions of *V. vulnificus* and *V. parahaemolyticus* infections followed an upward trend with increasing age group (both p<0.001), with the opposite pattern for *V. alginolyticus* (p<0.001), and no trend was observed for non-toxigenic *V. cholerae* infections (p=0.081) (Table 1).

Information on exposure was collected in two countries (Norway and Sweden) that constituted 239 cases of this study. The reported exposures were seawater/bathing (n=107, 44.8%), food/water poisoning (n=6, 2.5%), other (unspecified) (n=1, 0.4%) or unknown (n=125, 52.3%) (Table 1).

### Geographic distribution of vibriosis cases

The geographic distribution of the vibriosis cases differed between *Vibrio* species (Figure 2).

**Figure 2.**
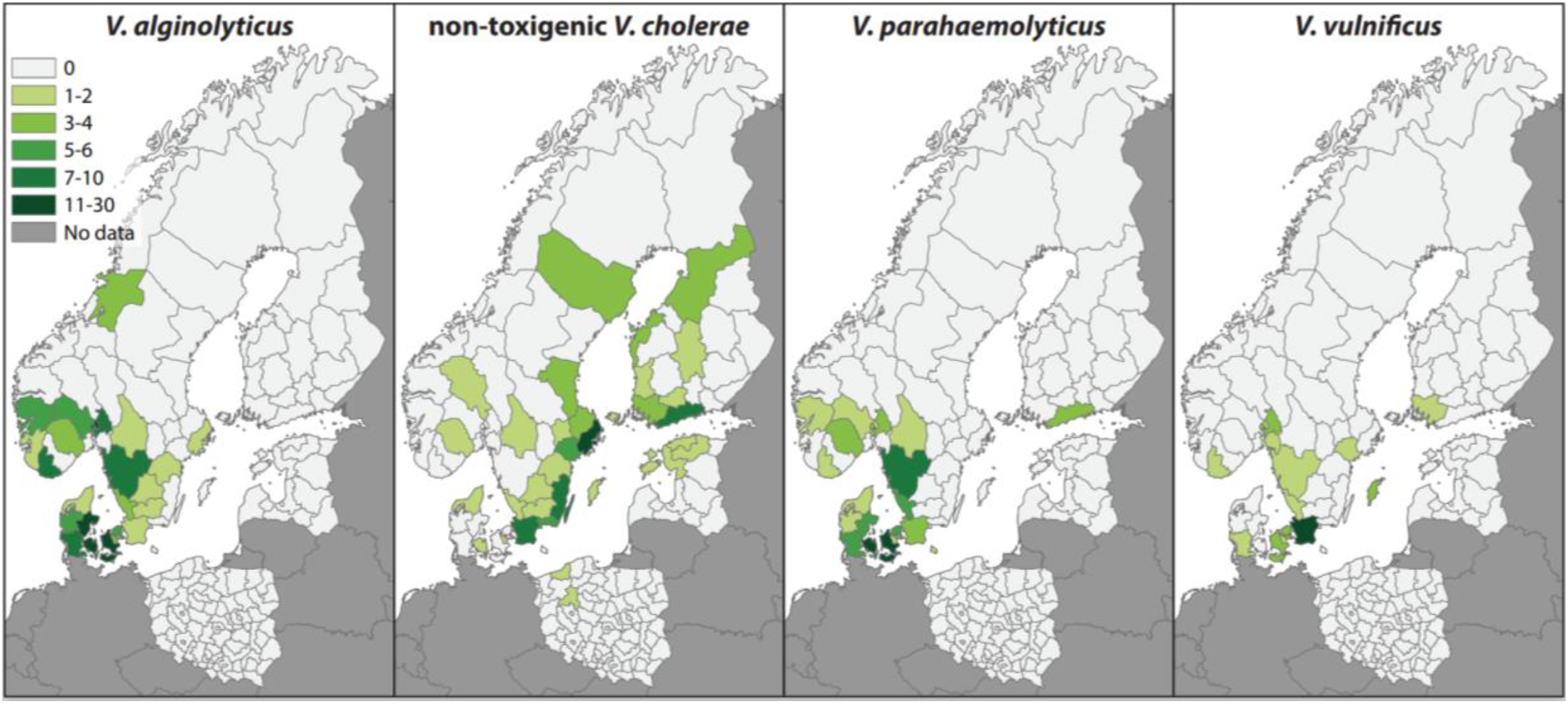
Geographical distribution (NUTS3 level) of vibriosis cases in relation to the identified species in the study countries in 2018. Note: Latvia reported no cases.

*V. alginolyticus* and *V. parahaemolyticus* infections were reported mainly from regions adjacent to the North Sea as well as around the connecting sounds between the Baltic and the North Sea: southern and western regions of Norway, all of Denmark and southwest coast of Sweden (Figure 2A and 2C). Non-toxigenic *V. cholerae* infections were almost exclusively reported from coastal regions of the Baltic Sea: the east coast of Sweden and regions in Finland, Poland, and Estonia (Figure 2B). *V. vulnificus* infections, similar to *V. alginolyticus* and *V. parahaemolyticus* infections, mainly occurred in the coastal regions around the connecting sounds between the Baltic and the North Sea, particularly Oslo fjord in Norway, southwest Sweden and eastern Denmark (Figure 2D).

### Severity of *Vibrio* infections

The proportion of severe VCV infections increased significantly with increasing age (p<0.001) and it differed by VCV (p<0.001) (Figure 3A and Table 2).

**Table 2.**
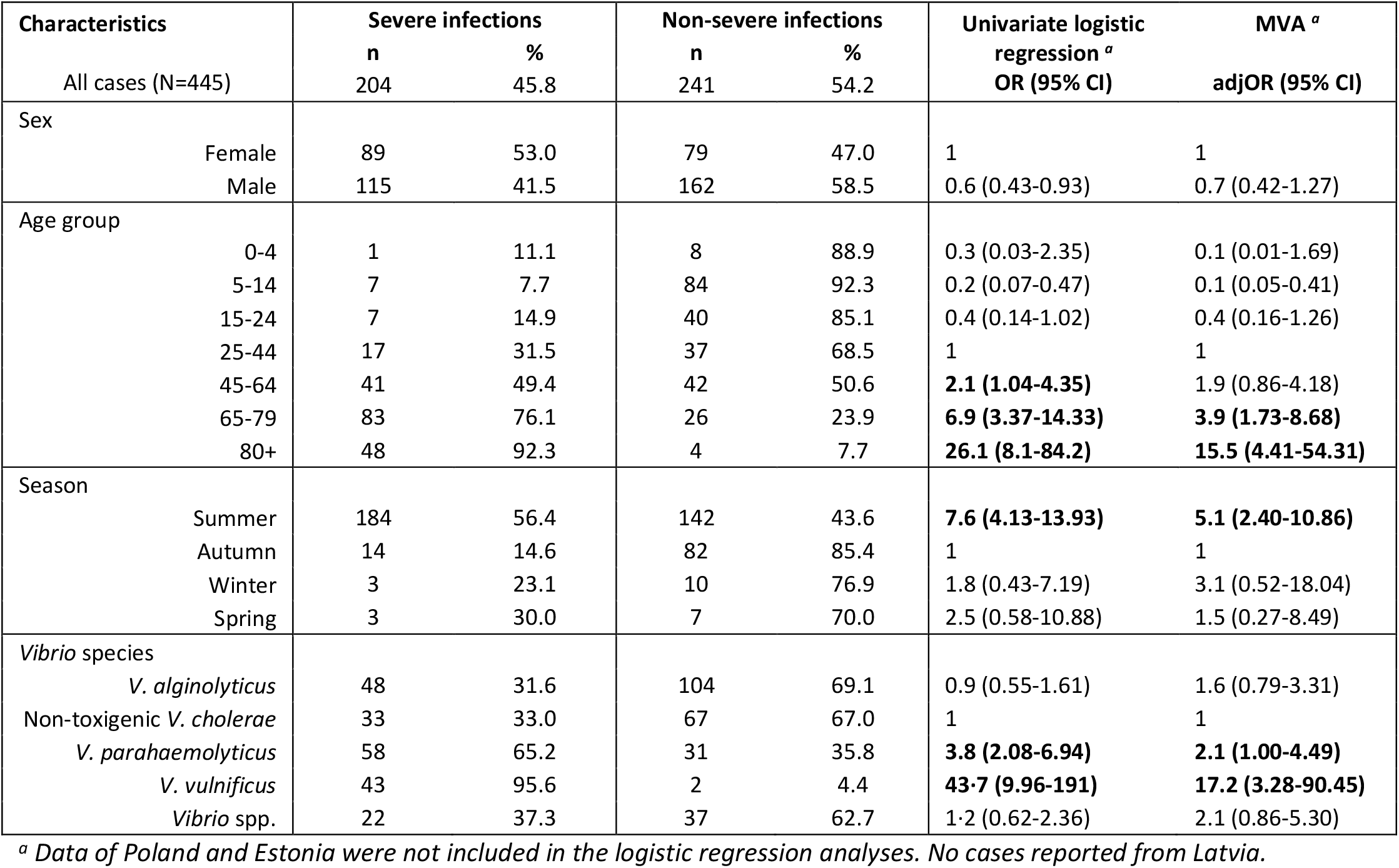
Predictors without and with adjustment of severe and non-severe vibriosis cases in the study countries, 2018.

**Figure 3.**
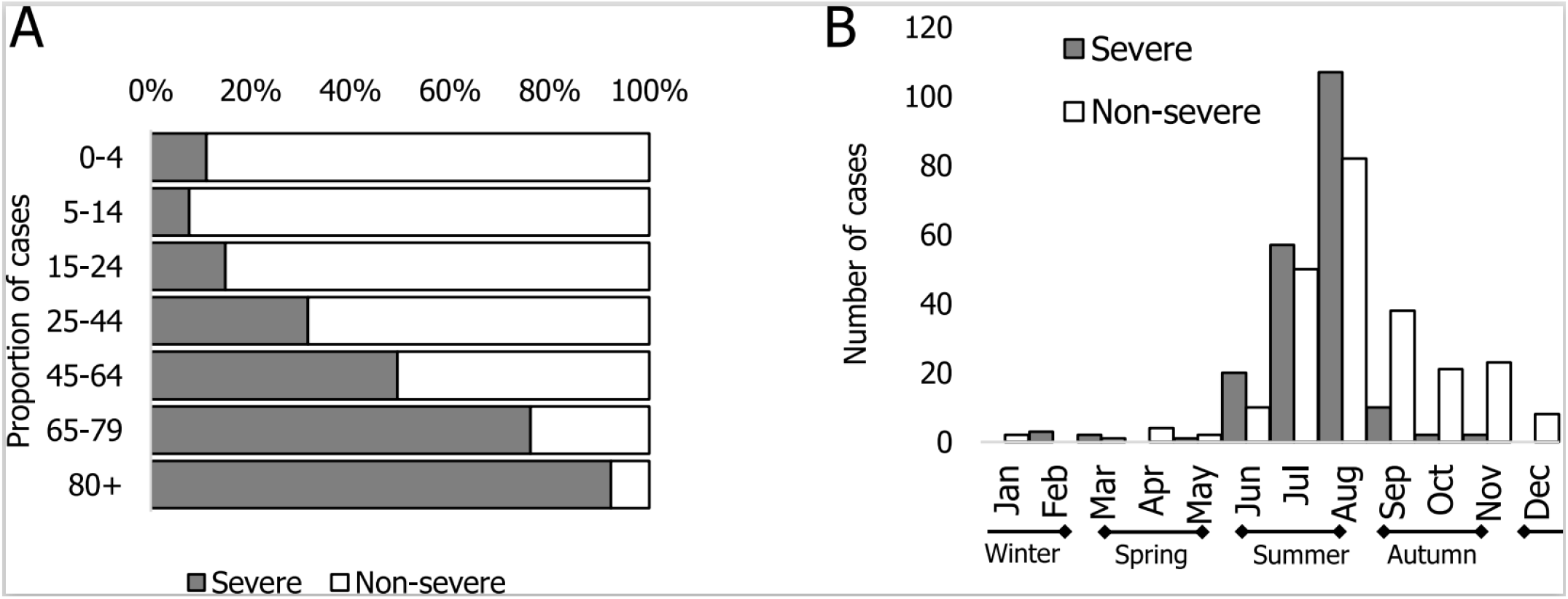
Severity of vibriosis cases. Proportion of cases per age group (A) and total numbers per months (B) in the study countries, 2018.

We observed the highest proportion of severe infections for *V. vulnificus* (95.6%) and *V. parahaemolyticus* (65.2%), while these were lower yet substantial for non-toxigenic *V. cholerae* (33.0%) and *V. alginolyticus* (31.6%) (Table 1 and 2). The exposure of these severe infections with non-toxigenic *V. cholerae* and *V. alginolyticus* was largely unknown (48% and 70%, respectively) or cases were exposed to seawater/bathing (48% and 25%, respectively). In terms of age, these infections were shifted slightly towards the younger age group. On the contrary, more than 70% of the severe *V. vulnificus* and *V. parahaemolyticus* infections occurred in the age groups 65-79 and 80+, while it was 58% and 41% of the severe *V. cholerae* and *V. alginolyticus* cases respectively that belonged to these age groups.

All VCV were more frequently reported in summer, when the majority of cases occurred (n=326, 73.3%; ranging per species from 63.8% to 97.8%) (Table 1). No difference in the seasonal distribution of vibriosis cases was observed between countries (Figure 3B, Table S2). According to our multivariable model, the likelihood of developing a severe infection was significantly increased among the elderly (65-79 years: adjOR=3.9; 95% CI: 1.7-8.7; 80+ years: adjOR=15.5; 95% CI: 4.4-54.3), for infections caused by *V. vulnificus* (adjOR=17.2; 95% CI: 3.3-90.5) or *V. parahaemolyticus* (adjOR=2.1; 95% CI: 1.0-4.5), as well as among infections occurring in summer (adjOR=5.1; 95% CI: 2.4-10.9) (Table 2).

### Microbiological and molecular investigations

We analysed whole genome sequences of 135 clinical *Vibrio* isolates isolated in 2018. Additionally, we included 16 available clinical isolates from travel related cases, 14 clinical isolates from 2014-2017 period and 13 Finnish environmental non-toxigenic *V. cholerae* isolates to investigate the genetic diversity of *Vibrio* in the study countries (Table S3).

#### Phylogenetic analysis

SNP analysis showed a high diversity of isolates for all species with several clusters of non-travel related cases (Figure 4 and 5, S1-S2).

Nine clusters with two or three cases each of non-toxigenic *V. cholerae* isolates (≤30 SNPs difference) were identified in Sweden (n=4), Sweden/Finland (n=4), and Finland (n=1) (Figure 4). Cases whose isolates clustered were sampled close in time (median 7 days; range 2-86 days) but detailed information on place of infection was not available.

**Figure 4.**
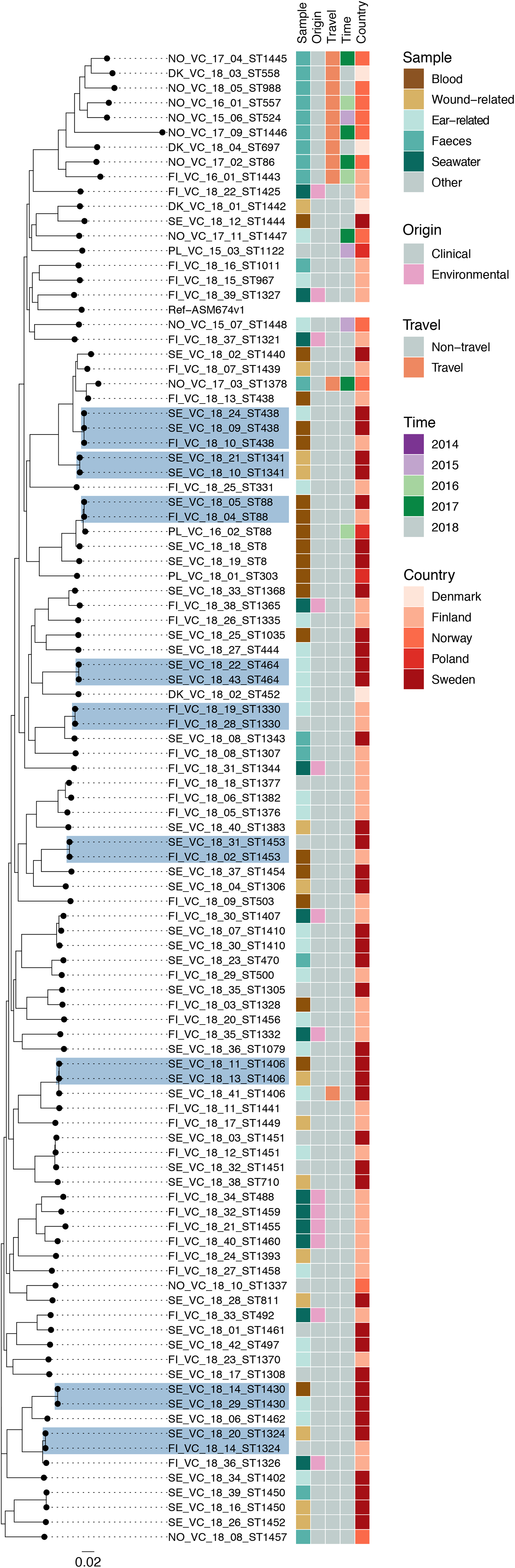
SNP based phylogeny of 100 non-toxigenic V. cholerae genomes from study countries. Nine V. cholerae clusters with ≤30 SNPs difference are shaded in steel blue. The non-toxigenic V. cholerae ASM674v1 sequence was used as reference. The scale bar indicates number of substitutions per site. Note: VC – non-toxigenic V. cholerae, DK – Denmark, FI – Finland, NO – Norway, SE – Sweden, ST – sequence type. The first number represents the isolation year and the second number denotes the isolate number.

**Figure 5.**
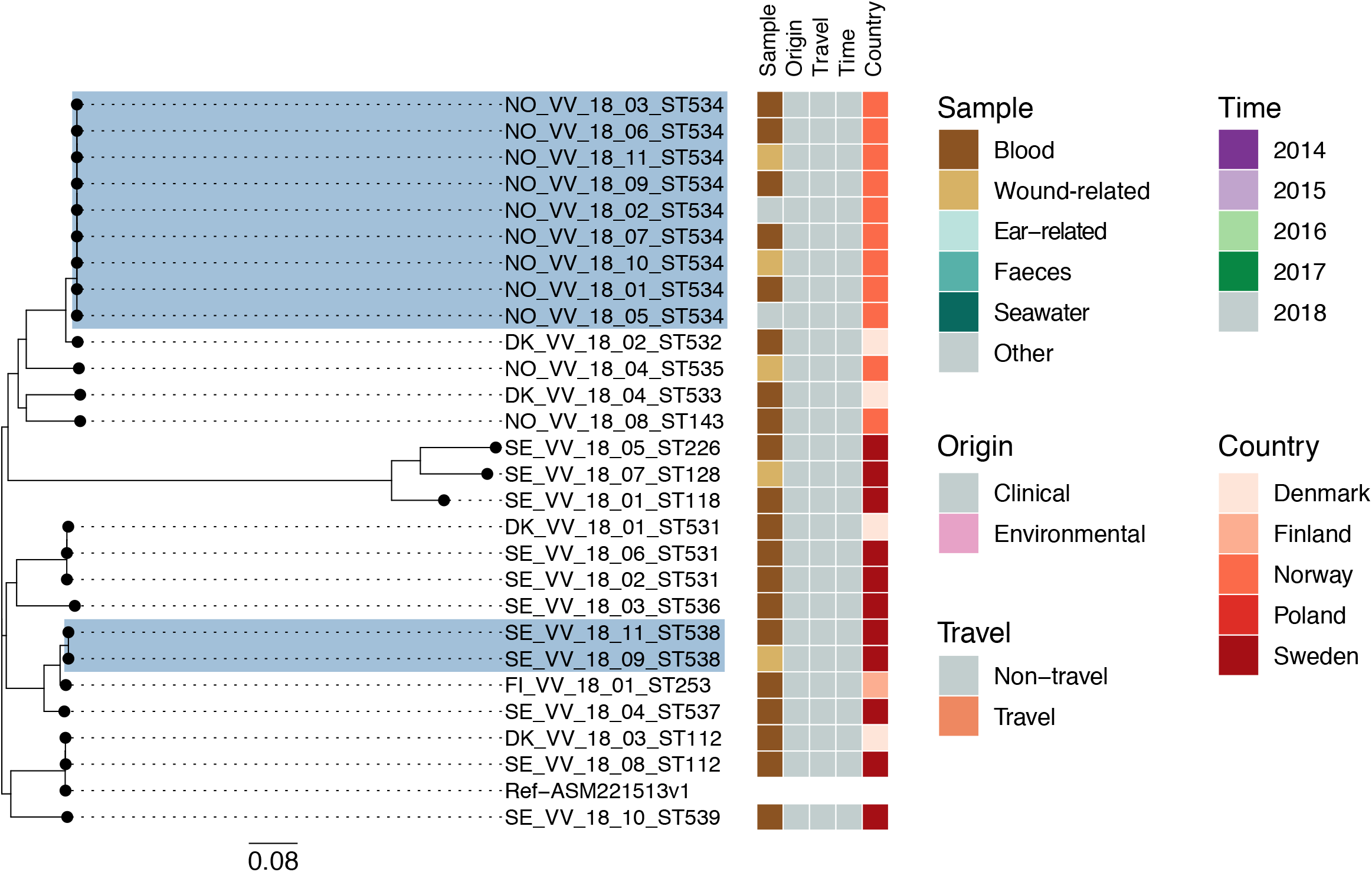
SNP based phylogeny of 27 V. vulnificus genomes from study countries. Two V. vulnificus clusters with ≤30 SNPs difference are shaded in steel blue. The V. vulnificus ASM221513v1 sequence was used as reference. The scale bar indicates the number of substitutions per site. Note: VV – V. vulnificus, DK – Denmark, FI – Finland, NO – Norway, SE – Sweden, ST – sequence type. The first number represents the isolation year and the second number denotes the isolate number.

Additionally, two clusters of *V. vulnificus* isolates with <10 SNPs difference were detected (Figure 5): one cluster with nine isolates in Norway, where the cases had been infected within 40 days and about 60 kilometres apart, and one cluster with two isolates in Sweden, where the cases had been infected 30 days and about 55 kilometres apart.

#### MLST analysis

Within the 178 isolates included in this study, 20 groups of isolates with the same ST were identified. Of these, ten groups were pairs of isolates from a single country (Sweden, Norway or Finland), three were pairs from two countries (Denmark/Sweden, n=1, or Sweden/Finland, n=2), six included 3-4 isolates each, and the largest group of nine *V. vulnificus* isolates (ST534) was detected in Norway (Figure 4 and 5, Table S4). Finally, a single *V. parahaemolyticus* isolate from Norway, found in a gastrointestinal infection in spring of 2014, was identified as the pandemic ST3 (Table S4, Figure S3).

## Discussion

Our study provides a detailed overview on the occurrence of vibriosis in the Nordic and Baltic Sea regions in 2018. In context of epidemiological and microbiological findings as well as conducted studies from 2014-2018 [11, 12], our results highlight the importance of vibriosis as a concern to public health in this geographic area. Even though the data have been collected using different systems, the study countries reported similar patterns in terms of population affected by sex and age-group distribution. Two-thirds of all vibriosis cases from 2014-2018 occurred in the years, 2014 and 2018, reported as two remarkably warm years in the literature [6, 11-14]. Moreover, though *V. vulnificus* infections are usually considered rare in this region [17], 45 such infections were detected in 2018 compared with the preceding years where eight (2014), none (2015), one (2016), and two (2017) *V. vulnificus* infections were identified. Interestingly, one *V. vulnificus* case occurred at about 60 degrees North latitude in Finland, which, to the best of our knowledge, is the highest latitude northward at which *V. vulnificus* has been reported. These findings underline a concern about the spread of this pathogen due to seawater warming [18].

It is well documented that vibriosis is more frequently reported in summer [2, 6, 11, 12, 18]. Our results from 2018 confirm this pattern with the majority of infections occurring in summer months. Additionally, in this study almost half of all infections reported in 2018 were categorised as severe infections that also mainly occurred during summer season. Mild ear infections may have long reporting delays up to months until a patient seeks medical care [19, 20] compared with rapidly developing severe blood or wound infections. We observed a similar pattern for the cases in this study, which could explain why reporting of mild vibriosis stretched more into autumn and winter and reporting of severe infections concentrated in summer months. More accurate information on the probable infection date would be needed to confirm this hypothesis. The likely source of infection was available for a subset of cases suggesting that the mode of transmission was mostly through seawater rather than through consumption of contaminated seafood.

The majority of vibriosis cases in the study countries were domestic and males were more frequently affected than females, consistent with other reports [12, 21]. Even though the majority of cases were present among adults, about a fifth of the detected cases were among children up to 14 years of age, who mostly had ear infections and mild vibriosis; severe infections on the other hand were found to be associated with increasing age. This is likely due to underlying conditions being overrepresented among elderly people. In addition to increasing age, we also found that being infected by *V. vulnificus* or *V. parahaemolyticus* was a risk factor for a more severe VCV infection likely due to the greater pathogenicity of these microorganisms [1, 2]. On the other hand, despite *V. cholerae* and *V. alginolyticus* predominantly causing mild infections, in our study, about one third of cases infected by these species were sampled from blood/serum or wounds. Thus, in absence of systematic data on hospitalisation and symptoms, these infections were also considered as severe. These cases were of a lower median age compared to *V. vulnificus* and *V. parahaemolyticus infections*, and the exposure was largely unknown with only some cases exposed to seawater/bathing. Given a substantial proportion of cases classified as severe, *V. cholerae* and *V. vulnificus* should therefore not be underestimated in vibriosis diagnosis, as was also pointed out previously [22].

We observed a geographic disparity in the distribution of VCV in the study countries. *V. alginolyticus* and *V. parahaemolyticus* infections concentrated in the coastal regions connecting the North Sea to the Baltic Sea, including the Danish Sounds, where *V. vulnificus* was mainly reported. Infections with non-toxigenic *V. cholerae* were mostly detected along the coasts of the Baltic Sea. This is in line with previous environmental detection of *Vibrio* species in different areas [2, 23-26] and reported clinical *V. vulnificus* infections from Germany [4]. Reasons for the geographic disparity could be related to differences in sea surface temperature and salinity, which represent major factors influencing *Vibrio* growth, and are continuously monitored in the European Centre for Disease Prevention and Control *Vibrio* suitability tool [27]. Additional factors, such as phytoplankton composition and nutrient presence in the water [23-25, 28], could have also played a role. Additional research studies on the water environment and presence of *Vibrio* in seafood could provide useful information on the ecological niches and geographical distribution of such bacteria particularly for species associated with a potentially severe clinical outcome. Our MLST analysis showed a genetic heterogeneity between clinical *Vibrio* spp. isolates, the majority of which belonged to new STs. SNP-based phylogenetic analysis revealed small clusters of *V. cholerae*, containing two to three isolates each, without a clear epidemiological link. The same *V. cholerae* strains detected in one or more countries might be due to common exposure to contaminated seafood or environmental spread of clones through e.g. sea currents [25], plastic pollutants [29], ship ballast water [30], and waterbirds [31]. The occurrence of two *V. vulnificus* clusters, one in Norway and one in Sweden, detected between 30-40 days apart and within an area of around 50-60 kilometres, highlights the possibility of emerging *V. vulnificus* clones that caused infections after seawater exposure during the exceptional warm summer in 2018. This was further supported by the epidemiological investigations of the first reported *V. vulnificus* waterborne outbreak after seawater exposure [32].

Some limitations apply to our investigation. There were differences in data sources and data availability between the study countries. Notification rates should therefore be compared carefully as vibriosis is not notifiable in all study countries or not for all species. Especially mild infections might have been reported with a delay and/or underreported. Conversely, in some cases a disease could have been misclassified as vibriosis when the identified *Vibrio* species were merely opportunistic microorganisms present at the site of infection. Case severity classification used in this analysis was not reported directly in any study country, but was inferred based on the sample type. Additionally, cases without known travel history were considered as non-travel related, which could have potentially led to an overestimation of vibriosis cases in the Baltic Sea region. Furthermore, the place of residence was used as proxy when place of infection was not available. Regarding the molecular findings, SNP analysis needs to be carefully evaluated since recombination is one of the major sources of genomic changes in *Vibrio*. Therefore, the removal of changes caused by recombination could have provided a better insight from the evolutionary perspective. Finally, laboratory methodology, capacity and priorities to diagnose and report VCV infections likely differed among the study countries.

During our investigation, we have performed a systematic and consistent analysis of epidemiological data from different countries, and we have combined it with the genomic analysis of strains from cases to achieve a comprehensive understanding of the occurrence of VCV infections in this affected region. Despite the low incidence, severe VCV infections are clinically costly [33] and climate changing predictions as well as population and socioeconomic projections for the upcoming years suggest that they are likely to increase in the future due to more favourable growth conditions for VCV [18, 34]. It is therefore of interest to detect and report the VCV infections in countries bordering the Baltic Sea and connecting regions to the North Sea to further monitor the situation, especially during summer heatwaves. Moreover, such surveillance would facilitate risk assessments and allow for targeted interventions, including risk communication to raise awareness between clinicians and populations at risk towards vibriosis. Thereby, countries without comprehensive surveillance could benefit from establishing or expanding dedicated surveillance systems to detect and prevent vibriosis cases. In particular, a shared sentinel system during summer months might be highly valuable.

## Data Availability

The dataset analysed in the study contains individual-level data from various central health registries or laboratory databases from participating countries. Only fully anonymised data (i.e. data that are neither directly nor potentially indirectly identifiable) are permitted to be shared publicly. Therefore, legal restrictions prevent the researchers from publicly sharing the dataset used in the study. However, external researchers are freely able to request access to linked data to each national public health institute as per normal procedure for conducting health research in respective countries in line with national and European regulations concerning data protection. Sequencing data of isolates are public available through the European Nucleotide Archive (ENA).

## Conflict of interest

None declared.

## Funding statement

The project has been financially supported by the European Programme for Public Health Microbiology Training (EUPHEM), ECDC. The funder had no role in study design, data collection and interpretation, or the decision to submit the work for publication. The views and opinions presented in this article are those of the authors and do not represent the opinion of the ECDC.

## Acknowledgements

The project has been financially supported by the European Programme for Public Health Microbiology Training (EUPHEM), ECDC and National Public Health Institutes of the authors’ countries. We would like to acknowledge Aftab Jasir, Silvia Herrera, Loredana Ingrosso and Aura Andreasen (EPIET/EUPHEM ECDC Fellowship Programme) for carefully revising this manuscript. Larisa Savrasova providing available information from Latvia and revision of the manuscript. We would like to thank Johanna Takkinen, Jan Semenza, Anders Dalsgaard, Yaovi Mahuton Gildas Hounmanou, Hans Blystad, Lotta Siira, Lamprini Veneti and Astrid Louise Wester for their scientific advice; Aino Kyyhkynen, Marja Veckström, Ingela Hedenström and Ina Patricia Haagensen for technical assistance in the laboratory. We thank the laboratories network from the study countries for providing VCV isolates and data.

## Contributors

EA, MR, DTL, ML collected the data at national level, jointly performed the epidemiological and microbiological analysis and prepared the manuscript. JAS carried out the bioinformatic analysis. TP contributed with the collection of environmental VCV isolates and interpretation of the analysis. TW and JR contributed in the analysis for the data and VCV isolates from Poland and Estonia. CJ advised during collecting and handling the clinical VCV isolates from Swedish laboratories and data interpretation. AH and KD collected epidemiological data in Sweden and Denmark, respectively. AH, MH, EMD and KD advised during epidemiological analysis and interpretation. AH, SS, KF and UN supervised the project at national level. UN supervised the project from the protocol development until the analysis and interpretation of results. All authors contributed to the manuscript, revised it and approved the final version.

## Notes

### Competing Interest Statement

The authors have declared no competing interest.

### Summary of Updates

Last sentence of the abstract available in the medRxiv search page is now harmonised with the abstract text in the archived PDF manuscript.

