## Supplementary material for "Epidemiological and microbiological investigation of the large increase of vibriosis in northern Europe in 2018"

### ***Table S1.*** *Characteristics of national surveillance systems of vibriosis in study countries, 2018*

| Country | Surveillance introduced | Data source | Reporting | Comprehensiveness | Design | Data reported by[1] | Data collection on all types of samples, regardless of infection severity[2] |
|---|---|---|---|---|---|---|---|
| Norway | No[3] | Laboratory database | Compulsory | Other[3, 4] | Passive | L | Yes |
| Sweden | Yes | National surveillance | Compulsory | Comprehensive | Passive | L, Ph, H | Yes |
| Denmark | No[5] | Laboratory database | Compulsory[5] | Other[5] | Passive | L | Yes |
| Finland | Yes | National surveillance | Compulsory | Comprehensive | Passive | L | Yes |
| Poland | Yes | National surveillance | Compulsory | Other[6] | Passive | L, Ph | Yes |
| Estonia | Yes | Laboratory database | Compulsory | Comprehensive | Passive | L | Yes |
| Latvia | No | - | - | - | - | - | - |

[1] *Data reported by: L – laboratories, Ph – physicians, H – hospitals.*

[2] *Severe infections (e.g. septicaemia, wound infections and necrotizing fasciitis) and non-severe infections (e.g. otitis, skin infections, gastroenteritis).*

[3] *Vibrio cholerae cases are routinely notified and isolates sent to the national reference laboratory at the national institute for the detection of cholera markers, wereas for non-cholerae Vibrio spp. this was the case for V. parahaemolyticus isolated from patients with gastrointestinal infections, only. Until 2019, other vibriosis were reported and isolates forwarded to the national institute in case of suspected outbreaks. Since 2019, vibriosis (irrespective of detected species) is a madatory notifiable disease in Norway.*

[4] *Data collected through nationwide laboratories survey for the year 2018.*

[5] *Laboratory findings are automatically logged into the Danish Microbiology Database (MiBA), including infections caused by all* Vibrio *species.*

[6] *Vibriosis cases were reported in the group "other bacterial foodborne infections" with only V. parahaemolyticus (ICD10 A05.3) specified as a separate vibriosis pathogen.*

### ***Table S2.*** *Summary of key characteristics of vibriosis cases in the study countries, 2018.*

| Characteristic | Countries | | | | | Total |
|---|---|---|---|---|---|---|
| | Denmark | Sweden | Norway | Finland | Poland and Estonia | |
| Reported cases, n [a] | 170 | 147 | 92 | 30 | 6 | 445 |
| Male-to-female ratio (cases) | 1.5 (101/69) | 2.2 (101/46) | 1.0 (47/45) | 4.0 (24/6) | 2.0 (4/2) | 1.6 (277/168) |
| Notification rate per 100,000 | 2.9 | 1.4 | 1.7 | 0.5 | ND | ND |
| Age, years | | | | | | |
| median | 53 | 58 | 42 | 39 | 34 | ND |
| range | 2-93 | 1-101 | 3-91 | 3-87 | 10-56 | 1-101 |
| Age group, n (%) | | | | | | |
| 0-4 | 2 (1.2%) | 3 (2%) | 2 (2.2%) | 2 (6.7%) | 0 (0%) | 9 (2%) |
| 5-14 | 34 (20%) | 24 (16.3%) | 24 (26.1%) | 7 (23.3%) | 2 (33.3%) | 91 (20.4%) |
| 15-24 | 20 (11.8%) | 14 (9.4%) | 10 (10.8%) | 2 (6.7%) | 1 (16.7%) | 47 (10.6%) |
| 25-44 | 20 (11.8%) | 16 (10.9%) | 13 (14.1%) | 5 (16.7%) | 0 (0%) | 54 (12.1%) |
| 45-64 | 28 (16.4%) | 31 (21.1%) | 18 (19.6%) | 4 (13.3%) | 2 (33.3%) | 83 (18.7%) |
| 65-79 | 49 (28.8%) | 35 (23.8%) | 18 (19.6%) | 6 (20%) | 1 (16.7%) | 109 (24.5%) |
| 80+ | 17 (10%) | 24 (16.3%) | 7 (7.6%) | 4 (13.3%) | 0 (0%) | 52 (11.7%) |
| Season, n (%) | | | | | | |
| summer | 134 (78.8%) | 109 (74.2%) | 57 (61.9%) | 22 (73.4%) | 4 (66.7%) | 326 (73.3%) |
| autumn | 29 (17.1%) | 29 (19.7%) | 31 (33.7%) | 6 (20%) | 1 (16.7%) | 96 (21.6%) |
| winter | 4 (2.3%) | 5 (3.4%) | 2 (2.2%) | 1 (3.3%) | 1 (16.6%) | 13 (2.9%) |
| spring | 3 (1.8%) | 4 (2.7%) | 2 (2.2%) | 1 (3.3%) | 0 (0%) | 10 (2.2%) |
| *Vibrio* species, n (%) | | | | | | |
| *V. alginolyticus* | 70 (41.2%) | 19 (12.9%) | 63 (68.5%) | 0 (0) | 0 (0) | 152 (34.2%) |
| Non-toxigenic *V. cholerae* | 3 (1.8%) | 64 (43.5%) | 2 (2.2%) | 26 (86.7%) | 5 (83.3%) | 100 (22.5%) |
| *V. parahaemolyticus* | 55 (32.4%) | 19 (12.9%) | 12 (13.0%) | 3 (10.0%) | 0 (0) | 89 (20.0%) |
| *V. vulnificus* | 16 (9.4%) | 19 (12.9%) | 9 (9.8%) | 1 (3.3%) | 0 (0) | 45 (10.1%) |
| *Vibrio* spp. | 26 (15.3%) | 26 (17.7%) | 6 (6.5%) | 0 (0) | 1 (16.7%) | 59 (13.3%) |

[a] *Data source: mandatory surveillance system (Sweden); nationwide surveying of public health microbiology laboratories (Norway); reporting of clinical sample analyses (Denmark, Finland, Poland, and Estonia). Latvia reported no cases. See material and methods for details on data acquisition. Note: ND – not determined.*

***Table S3.*** *Whole genome sequenced Vibrio isolates in the study countries, 2014-2018.*

| **Species** | **Country** | | | | | | | | | | | | |
|---|---|---|---|---|---|---|---|---|---|---|---|---|---|
| | **Norway** | | **Denmark** | | **Sweden** | | **Finland** | | | **Poland** | | **Total** | | |
| | **nT** | **T** | **nT** | **T** | **nT** | **T** | **nT** | **T** | **Env** | **nT** | **T** | **nT** | **T** | **Env** |
| *V. alginolyticus* | 16 (7) | 0 (1) | 0 | 0 | 6 | 1 | 0 | 0 | 0 | 0 | 0 | 22 (7) | 1 (1) | 0 |
| *Non- toxigenic V. cholerae* | 2 (2) | 1 (6) | 2 | 2 | 41 | 1 | 26 | 0 (1) | 13 | 1(2) | 0 | 72 (4) | 4 (7) | 13 |
| *V. parahaemolyticus* | 6 (3) | 2 (1) | 0 | 0 | 8 | 0 | 0 | 0 | 0 | 0 | 0 | 14 (3) | 2 (1) | 0 |
| *V. vulnificus* | 11 | 0 | 4 | 0 | 11 | 0 | 1 | 0 | 0 | 0 | 0 | 27 | 0 | 0 |
| **Total** | 35 (12) | 3 (8) | 6 | 2 | 66 | 2 | 27 | 0 (1) | 13 | 1(2) | 0 | 135 (14) | 7 (9) | 13 |

nT – non-travel related isolates or travel status unknown; T – travel related isolates; Env – environmental. Numbers indicate isolates of 2018 and in parenthesis isolates collected during 2014-2017.

***Table S4.*** *Summary table of assembly and FASTQ statistics for VCV isolates from study countries, 2014-2018.*

| Isolate name[1] | Sequence type | Accession no. | Assembly statistics | | | | FASTQ Statistics | | | |
|---|---|---|---|---|---|---|---|---|---|---|
| | | | Genome size | No. Contigs | Average size | N50 | No Reads | Coverage | Mean Size read | SD Read length |
| NO_VA_18_19 | 96 | ERS5890681 | 5421232 | 176 | 30802.45 | 315982 | 807569 | 236.415 | 146.374 | 14.7044 |
| NO_VA_18_06 | 138 | ERS5890687 | 5417902 | 190 | 28515.27 | 217049 | 948785 | 277.553 | 146.267 | 15.1111 |
| NO_VA_18_01 | 180 | ERS5890743 | 5407027 | 145 | 37289.84 | 276131 | 958833 | 283.302 | 147.733 | 11.0525 |
| SE_VA_18_07 | 181 | ERS5890679 | 5595925 | 107 | 52298.36 | 341854 | 3128519 | 883.332 | 141.174 | 21.1386 |
| NO_VA_18_13 | 182 | ERS5890690 | 5329433 | 158 | 33730.59 | 261597 | 826886 | 239.971 | 145.105 | 17.4573 |
| NO_VA_18_22 | 183 | ERS5890693 | 5374229 | 125 | 42993.83 | 301521 | 1215618 | 358.178 | 147.324 | 12.6567 |
| NO_VA_18_02 | 184 | ERS5890691 | 5262937 | 164 | 32091.08 | 236357 | 1009798 | 298.569 | 147.836 | 10.7947 |
| SE_VA_18_05 | 185 | ERS5890742 | 5294713 | 232 | 22822.04 | 335172 | 3332205 | 934.654 | 140.246 | 21.9543 |
| NO_VA_18_18 | 186 | ERS5890739 | 5161131 | 106 | 48689.92 | 474871 | 616064 | 182.035 | 147.74 | 11.4923 |
| NO_VA_14_12 | 187 | ERS5890686 | 5407567 | 164 | 32972.97 | 374289 | 555814 | 162.126 | 145.845 | 16.0321 |
| NO_VA_18_24 | 188 | ERS5890695 | 5156135 | 205 | 25151.88 | 243721 | 906604 | 263.139 | 145.124 | 17.2805 |
| NO_VA_17_03 | 189 | ERS5890688 | 5123679 | 145 | 35335.72 | 377119 | 668523 | 197.049 | 147.377 | 12.0064 |
| SE_VA_18_01 | 189 | ERS5890699 | 5209961 | 206 | 25291.07 | 306291 | 2342028 | 668.262 | 142.667 | 19.0984 |
| SE_VA_18_03 | 189 | ERS5890683 | 5245702 | 178 | 29470.24 | 306620 | 1991680 | 570.277 | 143.165 | 18.6206 |
| NO_VA_14_08 | 190 | ERS5890684 | 5344746 | 125 | 42757.97 | 285297 | 919872 | 268.354 | 145.865 | 15.9698 |
| NO_VA_18_21 | 190 | ERS5890698 | 5248406 | 113 | 46446.07 | 424596 | 859013 | 250.954 | 146.071 | 15.5318 |
| NO_VA_14_09 | 191 | ERS5890697 | 5126912 | 195 | 26291.86 | 184437 | 1063035 | 304.936 | 143.427 | 20.0946 |
| NO_VA_14_10 | 192 | ERS5890689 | 4856867 | 687 | 7069.68 | 12563 | 419714 | 109.1 | 129.97 | 31.0324 |
| NO_VA_16_14 | 193 | ERS5890741 | 5275994 | 185 | 28518.89 | 245908 | 1599715 | 450.432 | 140.785 | 23.0207 |
| SE_VA_18_02 | 194 | ERS5890678 | 5105536 | 150 | 34036.91 | 416418 | 2725118 | 786.1 | 144.232 | 16.6848 |
| NO_VA_18_16 | 195 | ERS5890685 | 5175692 | 190 | 27240.48 | 218225 | 381787 | 111.068 | 145.458 | 15.9461 |
| NO_VA_18_17 | 196 | ERS5890696 | 5199821 | 124 | 41934.04 | 486539 | 1030903 | 302.282 | 146.61 | 14.3043 |
| NO_VA_15_20 | 197 | ERS5890737 | 5221210 | 112 | 46617.95 | 461113 | 697234 | 204.871 | 146.917 | 13.571 |
| NO_VA_18_11 | 198 | ERS5890677 | 5371427 | 247 | 21746.67 | 262452 | 770781 | 224.259 | 145.475 | 16.5306 |
| SE_VA_18_06 | 199 | ERS5890740 | 5092698 | 148 | 34410.12 | 291124 | 2967297 | 846.333 | 142.61 | 19.1673 |
| NO_VA_18_15 | 200 | ERS5890682 | 5276788 | 213 | 24773.65 | 316620 | 2126896 | 594.344 | 139.721 | 24.7359 |
| NO_VA_18_05 | 201 | ERS5890694 | 5342558 | 191 | 27971.51 | 197410 | 2610356 | 716.865 | 137.312 | 27.1104 |
| NO_VA_18_07 | 202 | ERS5890680 | 5268605 | 123 | 42834.19 | 267069 | 1261658 | 363.711 | 144.14 | 19.021 |
| NO_VA_18_23 | 202 | ERS5890692 | 5204394 | 124 | 41970.92 | 368527 | 823506 | 241.841 | 146.836 | 14.0369 |
| SE_VA_18_04 | 203 | ERS5890738 | 5075931 | 187 | 27144.02 | 392950 | 823252 | 245.081 | 148.849 | 7.12042 |
| NO_VA_14_04 | 204 | ERS5890676 | 5179946 | 165 | 31393.61 | 446902 | 1318903 | 380.827 | 144.373 | 17.7441 |
| SE_VC_18_18 | 8 | ERS5890780 | 4085592 | 165 | 24761.16 | 177775 | 2515588 | 707.471 | 140.618 | 21.6816 |
| SE_VC_18_19 | 8 | ERS5890762 | 3999033 | 122 | 32778.96 | 247924 | 2724814 | 769.103 | 141.129 | 21.5285 |
| NO_VC_17_02 | 86 | ERS5890648 | 3994385 | 228 | 17519.23 | 86125 | 1356159 | 380.053 | 140.121 | 24.0678 |
| FI_VC_18_04 | 88 | ERS5890649 | 4123998 | 410 | 10058.53 | 148081 | 1197898 | 351.059 | 146.531 | 14.0905 |
| PL_VC_16_02 | 88 | ERS5890660 | 3992736 | 139 | 28724.72 | 155401 | 1091402 | 317.057 | 145.252 | 17.2512 |
| SE_VC_18_05 | 88 | ERS5890663 | 3966014 | 103 | 38504.99 | 155419 | 2563988 | 717.34 | 139.888 | 22.6501 |

| | | | | | | | | | | |
|---|---|---|---|---|---|---|---|---|---|---|
| PL_VC_18_01 | 303 | ERS5890665 | 3987824 | 158 | 25239.39 | 246279 | 903757 | 264.063 | 146.092 | 15.5191 |
| FI_VC_18_25 | 331 | ERS5890668 | 4060283 | 228 | 17808.26 | 253959 | 2069404 | 606.635 | 146.573 | 14.0851 |
| FI_VC_18_10 | 438 | ERS5890645 | 4014291 | 281 | 14285.73 | 249848 | 2111542 | 619.075 | 146.593 | 14.0151 |
| FI_VC_18_13 | 438 | ERS5890640 | 4142630 | 312 | 13277.66 | 211126 | 1681522 | 492.875 | 146.556 | 14.0625 |
| SE_VC_18_09 | 438 | ERS5890744 | 3952491 | 144 | 27447.85 | 249848 | 1361757 | 398.922 | 146.473 | 12.8524 |
| SE_VC_18_24 | 438 | ERS5890703 | 3964416 | 199 | 19921.69 | 210781 | 3484277 | 990.524 | 142.142 | 19.7046 |
| SE_VC_18_27 | 444 | ERS5890734 | 3969652 | 204 | 19459.08 | 302560 | 2240015 | 636.282 | 142.026 | 20.197 |
| DK_VC_18_02 | 452 | ERS5890673 | 4065772 | 132 | 30801.3 | 128504 | 1368935 | 402.871 | 147.148 | 12.8087 |
| SE_VC_18_22 | 464 | ERS5890707 | 3946328 | 93 | 42433.63 | 271865 | 1410684 | 415.327 | 147.208 | 11.5934 |
| SE_VC_18_43 | 464 | ERS5890630 | 3948644 | 71 | 55614.7 | 572877 | 2958774 | 837.746 | 141.57 | 20.7317 |
| SE_VC_18_23 | 470 | ERS5890735 | 4023206 | 120 | 33526.72 | 217285 | 2355634 | 666.511 | 141.472 | 20.9195 |
| FI_VC_18_34 | 488 | ERS5890652 | 4460675 | 660 | 6758.6 | 210058 | 1520824 | 445.637 | 146.512 | 14.2519 |
| FI_VC_18_33 | 492 | ERS5890639 | 4120663 | 466 | 8842.62 | 237533 | 1778598 | 520.152 | 146.225 | 14.7686 |
| SE_VC_18_42 | 497 | ERS5890709 | 4044664 | 220 | 18384.84 | 184342 | 993932 | 295.96 | 148.884 | 7.38207 |
| FI_VC_18_29 | 500 | ERS5890643 | 4048074 | 317 | 12769.95 | 137653 | 1958806 | 574.459 | 146.635 | 13.9492 |
| FI_VC_18_09 | 503 | ERS5890662 | 4120619 | 322 | 12796.95 | 345276 | 2218474 | 645.657 | 145.518 | 16.1659 |
| NO_VC_15_06 | 524 | ERS5890755 | 4093649 | 191 | 21432.72 | 185142 | 563896 | 166.854 | 147.947 | 10.7875 |
| NO_VC_16_01 | 557 | ERS5890774 | 3925971 | 193 | 20341.82 | 146958 | 1224335 | 334.402 | 136.565 | 27.5007 |
| DK_VC_18_03 | 558 | ERS5890672 | 3988972 | 234 | 17046.89 | 141847 | 1514790 | 424.313 | 140.057 | 23.8934 |
| DK_VC_18_04 | 697 | ERS5890670 | 3884611 | 346 | 11227.2 | 111596 | 672384 | 196.088 | 145.815 | 15.5662 |
| SE_VC_18_38 | 710 | ERS5890765 | 4053433 | 254 | 15958.4 | 165378 | 2696100 | 774.056 | 143.551 | 17.9423 |
| SE_VC_18_28 | 811 | ERS5890764 | 4324690 | 289 | 14964.33 | 181503 | 2082543 | 593.734 | 142.55 | 19.347 |
| FI_VC_18_15 | 967 | ERS5890772 | 4254710 | 253 | 16817.04 | 150709 | 2078129 | 608.467 | 146.398 | 14.3373 |
| NO_VC_18_05 | 988 | ERS5890778 | 3951913 | 229 | 17257.26 | 152892 | 688787 | 200.279 | 145.385 | 16.69 |
| FI_VC_18_16 | 1011 | ERS5890627 | 4092488 | 209 | 19581.28 | 238852 | 1056143 | 308.994 | 146.284 | 14.5634 |
| SE_VC_18_25 | 1035 | ERS5890783 | 4048058 | 141 | 28709.63 | 217348 | 2578716 | 733.379 | 142.199 | 19.839 |
| SE_VC_18_36 | 1079 | ERS5890708 | 4062026 | 276 | 14717.49 | 256663 | 2256469 | 649.969 | 144.023 | 17.2764 |
| PL_VC_15_03 | 1122 | ERS5890769 | 4128666 | 123 | 33566.39 | 235089 | 700622 | 207.432 | 148.034 | 10.5464 |
| SE_VC_18_35 | 1305 | ERS5890752 | 4073088 | 270 | 15085.51 | 142229 | 1487203 | 432.762 | 145.495 | 15.1259 |
| SE_VC_18_04 | 1306 | ERS5890773 | 4013120 | 138 | 29080.58 | 270481 | 2329456 | 651.594 | 139.86 | 23.1752 |
| FI_VC_18_08 | 1307 | ERS5890756 | 4184733 | 349 | 11990.64 | 248241 | 2044865 | 598.571 | 146.36 | 14.5095 |
| SE_VC_18_17 | 1308 | ERS5890747 | 4051967 | 345 | 11744.83 | 311295 | 2118976 | 603.946 | 142.509 | 19.3805 |
| FI_VC_18_37 | 1321 | ERS5890664 | 5496910 | 3445 | 1595.62 | 3884 | 1769208 | 517.704 | 146.309 | 14.5189 |
| FI_VC_18_14 | 1324 | ERS5890651 | 4083111 | 380 | 10745.03 | 253365 | 1711347 | 501.994 | 146.666 | 13.8321 |
| SE_VC_18_20 | 1324 | ERS5890701 | 4025728 | 163 | 24697.72 | 203908 | 2562805 | 727.303 | 141.896 | 20.2302 |
| FI_VC_18_36 | 1326 | ERS5890736 | 5875531 | 5050 | 1163.47 | 2059 | 2834886 | 830.26 | 146.436 | 14.2883 |
| FI_VC_18_39 | 1327 | ERS5890777 | 6179601 | 4604 | 1342.22 | 2523 | 1921632 | 563.465 | 146.611 | 13.9655 |
| FI_VC_18_03 | 1328 | ERS5890771 | 4209040 | 336 | 12526.9 | 128998 | 944340 | 275.039 | 145.625 | 15.7838 |
| FI_VC_18_19 | 1330 | ERS5890657 | 4398920 | 556 | 7911.73 | 170863 | 1489878 | 435.609 | 146.189 | 14.8914 |
| FI_VC_18_28 | 1330 | ERS5890647 | 4093907 | 279 | 14673.5 | 181377 | 2067901 | 605.784 | 146.473 | 14.1882 |
| FI_VC_18_35 | 1332 | ERS5890656 | 4073160 | 335 | 12158.69 | 307485 | 2254360 | 661.036 | 146.613 | 13.9506 |
| FI_VC_18_26 | 1335 | ERS5890674 | 4057346 | 290 | 13990.85 | 198903 | 1605322 | 470.562 | 146.563 | 14.0913 |
| NO_VC_18_10 | 1337 | ERS5890761 | 4201468 | 400 | 10503.67 | 179016 | 1536627 | 437.171 | 142.25 | 21.4665 |
| SE_VC_18_10 | 1341 | ERS5890776 | 3972658 | 153 | 25965.08 | 240903 | 2670166 | 765.723 | 143.385 | 18.1254 |
| SE_VC_18_21 | 1341 | ERS5890750 | 3972808 | 154 | 25797.45 | 240771 | 2664167 | 758.575 | 142.366 | 19.6193 |
| SE_VC_18_08 | 1343 | ERS5890775 | 4579021 | 464 | 9868.58 | 55554 | 2399122 | 683.934 | 142.538 | 19.4236 |
| FI_VC_18_31 | 1344 | ERS5890650 | 4060116 | 411 | 9878.63 | 200339 | 2338586 | 684.911 | 146.437 | 14.3523 |
| FI_VC_18_38 | 1365 | ERS5890784 | 6036664 | 5200 | 1160.9 | 2041 | 2654973 | 778.285 | 146.571 | 14.0229 |
| SE_VC_18_33 | 1368 | ERS5890706 | 4032620 | 152 | 26530.39 | 334555 | 2184891 | 624.574 | 142.93 | 18.9731 |
| FI_VC_18_23 | 1370 | ERS5890641 | 4264238 | 535 | 7970.54 | 259073 | 1847148 | 539.58 | 146.058 | 15.0553 |
| FI_VC_18_05 | 1376 | ERS5890655 | 4257789 | 435 | 9788.02 | 252415 | 1169773 | 341.342 | 145.901 | 15.3938 |
| FI_VC_18_18 | 1377 | ERS5890751 | 4240020 | 617 | 6871.99 | 202624 | 1413805 | 412.443 | 145.863 | 15.4104 |
| NO_VC_17_03 | 1378 | ERS5890754 | 3953247 | 142 | 27839.77 | 234977 | 606394 | 177.859 | 146.653 | 14.0592 |
| FI_VC_18_06 | 1382 | ERS5890646 | 4213066 | 503 | 8375.88 | 180126 | 1476545 | 432.998 | 146.625 | 13.9515 |
| SE_VC_18_40 | 1383 | ERS5890702 | 3993134 | 172 | 23215.9 | 185950 | 3204566 | 906.099 | 141.376 | 20.8627 |
| FI_VC_18_24 | 1393 | ERS5890635 | 4253855 | 429 | 9915.75 | 346686 | 1804724 | 528.006 | 146.284 | 14.6164 |
| SE_VC_18_34 | 1402 | ERS5890704 | 4091017 | 195 | 20979.57 | 190389 | 3289550 | 934.944 | 142.108 | 19.8586 |
| SE_VC_18_11 | 1406 | ERS5890745 | 4082959 | 230 | 17752 | 157007 | 2807358 | 805.358 | 143.437 | 18.033 |
| SE_VC_18_13 | 1406 | ERS5890766 | 4110006 | 252 | 16309.55 | 157007 | 2094804 | 598.724 | 142.907 | 19.0521 |
| SE_VC_18_41 | 1406 | ERS5890759 | 4094914 | 227 | 18039.27 | 157632 | 3005353 | 856.381 | 142.476 | 19.4825 |
| FI_VC_18_30 | 1407 | ERS5890661 | 4115955 | 391 | 10526.74 | 182989 | 2267493 | 664.326 | 146.489 | 14.2163 |
| SE_VC_18_07 | 1410 | ERS5890785 | 3997947 | 151 | 26476.47 | 200755 | 2355879 | 663.645 | 140.849 | 21.707 |
| SE_VC_18_30 | 1410 | ERS5890770 | 4048778 | 142 | 28512.52 | 200792 | 2657256 | 754.751 | 142.017 | 20.1819 |

| | | | | | | | | | | |
|---|---|---|---|---|---|---|---|---|---|---|
| FI_VC_18_22 | 1425 | ERS5890667 | 4207202 | 422 | 9969.67 | 195320 | 1471245 | 431.388 | 146.606 | 13.9786 |
| SE_VC_18_14 | 1430 | ERS5890705 | 4084169 | 277 | 14744.29 | 338961 | 2361486 | 676.297 | 143.193 | 18.515 |
| SE_VC_18_29 | 1430 | ERS5890753 | 4121961 | 289 | 14262.84 | 339029 | 2538178 | 720.615 | 141.955 | 20.417 |
| FI_VC_18_07 | 1439 | ERS5890638 | 4097939 | 353 | 11608.89 | 246500 | 1485128 | 435.006 | 146.454 | 14.3185 |
| SE_VC_18_02 | 1440 | ERS5890760 | 4027128 | 151 | 26669.72 | 246593 | 1871523 | 531.819 | 142.082 | 20.1539 |
| FI_VC_18_11 | 1441 | ERS5890636 | 4589003 | 773 | 5936.61 | 92152 | 1367795 | 400.869 | 146.539 | 14.1278 |
| DK_VC_18_01 | 1442 | ERS5890758 | 4105070 | 232 | 17694.27 | 90271 | 2427758 | 627.209 | 129.175 | 30.5484 |
| FI_VC_16_01 | 1443 | ERS5890710 | 4093432 | 374 | 10945.01 | 134899 | 1781910 | 522.741 | 146.68 | 13.8599 |
| SE_VC_18_12 | 1444 | ERS5890763 | 4070653 | 190 | 21424.49 | 171342 | 2933153 | 840.835 | 143.333 | 18.1592 |
| NO_VC_17_04 | 1445 | ERS5890669 | 4020954 | 199 | 20205.8 | 123186 | 520956 | 152.706 | 146.564 | 14.3174 |
| NO_VC_17_09 | 1446 | ERS5890659 | 3957109 | 227 | 17432.2 | 158315 | 577436 | 169.657 | 146.906 | 13.5845 |
| NO_VC_17_11 | 1447 | ERS5890757 | 3996588 | 175 | 22837.65 | 160525 | 783253 | 221.242 | 141.233 | 22.9442 |
| NO_VC_15_07 | 1448 | ERS5890628 | 4002234 | 170 | 23542.55 | 317992 | 431555 | 127.458 | 147.672 | 11.6002 |
| FI_VC_18_17 | 1449 | ERS5890700 | 4193491 | 524 | 8002.85 | 148010 | 2253099 | 661.224 | 146.737 | 13.6388 |
| SE_VC_18_16 | 1450 | ERS5890749 | 3925286 | 150 | 26168.57 | 199683 | 2192903 | 613.279 | 139.833 | 22.8064 |
| SE_VC_18_39 | 1450 | ERS5890782 | 3939055 | 172 | 22901.48 | 277126 | 1880124 | 538.351 | 143.169 | 18.6115 |
| FI_VC_18_12 | 1451 | ERS5890658 | 4199534 | 428 | 9812 | 131721 | 1859128 | 544.729 | 146.501 | 14.1309 |
| SE_VC_18_03 | 1451 | ERS5890746 | 4051940 | 184 | 22021.41 | 141073 | 1795077 | 511.278 | 142.411 | 19.9432 |
| SE_VC_18_32 | 1451 | ERS5890767 | 4043476 | 208 | 19439.79 | 132959 | 2272298 | 654.215 | 143.954 | 17.2797 |
| SE_VC_18_26 | 1452 | ERS5890629 | 4056334 | 150 | 27042.23 | 247076 | 3166412 | 897.225 | 141.678 | 20.5119 |
| FI_VC_18_02 | 1453 | ERS5890671 | 4295099 | 551 | 7795.1 | 258752 | 1745630 | 510.497 | 146.222 | 14.862 |
| SE_VC_18_31 | 1453 | ERS5890748 | 4083125 | 275 | 14847.73 | 259159 | 2191611 | 622.222 | 141.955 | 20.5207 |
| SE_VC_18_37 | 1454 | ERS5890786 | 4006908 | 173 | 23161.32 | 289893 | 1748603 | 498.275 | 142.478 | 19.6693 |
| FI_VC_18_21 | 1455 | ERS5890642 | 4217506 | 359 | 11747.93 | 299665 | 2016412 | 589.018 | 146.056 | 15.0806 |
| FI_VC_18_20 | 1456 | ERS5890666 | 4398960 | 503 | 8745.45 | 157740 | 1612774 | 473.043 | 146.655 | 13.8423 |
| NO_VC_18_08 | 1457 | ERS5890779 | 3922880 | 115 | 34112 | 248195 | 827482 | 243.432 | 147.092 | 13.1727 |
| FI_VC_18_27 | 1458 | ERS5890637 | 4096933 | 320 | 12802.92 | 276778 | 1773560 | 517.5 | 145.893 | 15.3816 |
| FI_VC_18_32 | 1459 | ERS5890654 | 4211988 | 350 | 12034.25 | 261943 | 2083644 | 611.023 | 146.624 | 13.9168 |
| FI_VC_18_40 | 1460 | ERS5890653 | 4253466 | 438 | 9711.11 | 514583 | 1636199 | 480.087 | 146.708 | 13.7508 |
| SE_VC_18_01 | 1461 | ERS5890781 | 4041774 | 224 | 18043.63 | 186142 | 2440169 | 692.796 | 141.957 | 20.193 |
| SE_VC_18_06 | 1462 | ERS5890768 | 4013885 | 230 | 17451.67 | 339329 | 2498845 | 686.51 | 137.365 | 25.9253 |
| NO_VP_14_01 | 3 | ERS5890798 | 5076372 | 127 | 39971.43 | 312643 | 1367888 | 398.367 | 145.614 | 15.5745 |
| NO_VP_18_11 | 77 | ERS5890719 | 4969676 | 94 | 52868.89 | 616547 | 614413 | 182.127 | 148.212 | 10.1077 |
| NO_VP_18_12 | 479 | ERS5890794 | 5358645 | 111 | 48276.08 | 515147 | 1778877 | 516.371 | 145.139 | 16.8569 |
| NO_VP_18_05 | 483 | ERS5890714 | 5340986 | 117 | 45649.45 | 361944 | 1000709 | 292.763 | 146.278 | 15.1041 |
| NO_VP_18_08 | 766 | ERS5890716 | 5051489 | 145 | 34837.86 | 331288 | 1562655 | 437.12 | 139.864 | 24.4253 |
| NO_VP_18_10 | 767 | ERS5890796 | 5162189 | 109 | 47359.53 | 327574 | 709200 | 208.573 | 147.048 | 13.3597 |
| SE_VP_18_04 | 1081 | ERS5890715 | 5086308 | 121 | 42035.6 | 455774 | 2697355 | 768.395 | 142.435 | 19.5158 |
| SE_VP_18_08 | 1346 | ERS5890792 | 5121318 | 118 | 43401 | 490626 | 2023103 | 577.674 | 142.769 | 19.1404 |
| NO_VP_18_02 | 1372 | ERS5890795 | 5026566 | 151 | 33288.52 | 193022 | 1780020 | 505.598 | 142.02 | 21.8689 |
| SE_VP_18_06 | 1815 | ERS5890791 | 5117517 | 75 | 68233.56 | 518931 | 3144612 | 896.21 | 142.499 | 19.2932 |
| NO_VP_17_07 | 2240 | ERS5890793 | 5266622 | 121 | 43525.8 | 469834 | 769824 | 222.389 | 144.442 | 18.5066 |
| NO_VP_18_09 | 2414 | ERS5890720 | 5289803 | 133 | 39772.95 | 390310 | 921920 | 269.202 | 146.001 | 15.8292 |
| SE_VP_18_01 | 2415 | ERS5890711 | 5019031 | 137 | 36635.26 | 329338 | 3343826 | 950.288 | 142.096 | 19.7324 |
| SE_VP_18_05 | 2416 | ERS5890713 | 5083570 | 158 | 32174.49 | 472036 | 2206774 | 632.948 | 143.41 | 18.2185 |
| NO_VP_18_03 | 2417 | ERS5890722 | 5164453 | 116 | 44521.15 | 539449 | 752819 | 220.308 | 146.322 | 14.9306 |
| SE_VP_18_02 | 2418 | ERS5890718 | 4926869 | 145 | 33978.41 | 326466 | 2821288 | 797.206 | 141.284 | 20.9407 |
| NO_VP_16_06 | 2419 | ERS5890797 | 5005092 | 152 | 32928.24 | 218250 | 2223756 | 585.281 | 131.597 | 30.951 |
| SE_VP_18_07 | 2420 | ERS5890721 | 5186572 | 118 | 43954 | 478064 | 2694201 | 762.377 | 141.485 | 20.781 |
| NO_VP_14_04 | 2421 | ERS5890717 | 5031640 | 110 | 45742.18 | 388862 | 711634 | 208.409 | 146.43 | 14.7628 |
| SE_VP_18_03 | 2423 | ERS5890712 | 5165336 | 73 | 70758.03 | 535832 | 2773380 | 784.773 | 141.483 | 20.8319 |
| DK_VV_18_03 | 112 | ERS5890803 | 4945660 | 302 | 16376.36 | 46709 | 1508698 | 398.522 | 132.075 | 29.4754 |
| SE_VV_18_08 | 112 | ERS5890724 | 5005813 | 313 | 15993.01 | 70053 | 2536020 | 725.399 | 143.019 | 18.5646 |
| SE_VV_18_01 | 118 | ERS5890631 | 5185854 | 352 | 14732.54 | 89585 | 2824385 | 807.726 | 142.991 | 18.629 |
| SE_VV_18_07 | 128 | ERS5890634 | 4828044 | 132 | 36576.09 | 192800 | 2513279 | 710.648 | 141.379 | 20.8896 |
| NO_VV_18_08 | 143 | ERS5890809 | 4984941 | 139 | 35862.88 | 220890 | 1090328 | 320.611 | 147.025 | 12.6556 |
| SE_VV_18_05 | 226 | ERS5890731 | 5047035 | 285 | 17708.89 | 120897 | 2592245 | 739.257 | 142.59 | 19.3631 |
| FI_VV_18_01 | 253 | ERS5890802 | 5104589 | 500 | 10209.18 | 205757 | 2029390 | 594.384 | 146.444 | 14.1642 |
| DK_VV_18_01 | 531 | ERS5890799 | 4953816 | 398 | 12446.77 | 145719 | 2215490 | 649.441 | 146.568 | 13.5376 |
| SE_VV_18_02 | 531 | ERS5890726 | 4966619 | 312 | 15918.65 | 131423 | 1987179 | 577.929 | 145.414 | 15.2668 |
| SE_VV_18_06 | 531 | ERS5890729 | 5029697 | 258 | 19494.95 | 131598 | 2585462 | 714.496 | 138.176 | 25.4798 |
| DK_VV_18_02 | 532 | ERS5890675 | 4980500 | 175 | 28460 | 161862 | 1613570 | 420.739 | 130.375 | 30.6238 |
| DK_VV_18_04 | 533 | ERS5890804 | 4929738 | 170 | 28998.46 | 103294 | 2046465 | 537.787 | 131.394 | 30.1469 |

| | | | | | | | | | | |
|---|---|---|---|---|---|---|---|---|---|---|
| NO_VV_18_06 | 534 | ERS5890806 | 4978003 | 165 | 30169.72 | 508086 | 795812 | 234.211 | 147.152 | 12.542 |
| NO_VV_18_07 | 534 | ERS5890807 | 4930202 | 151 | 32650.34 | 450798 | 884153 | 260.226 | 147.161 | 12.5734 |
| NO_VV_18_09 | 534 | ERS5890730 | 4930278 | 182 | 27089.44 | 380496 | 592833 | 174.728 | 147.367 | 12.2702 |
| NO_VV_18_10 | 534 | ERS5890732 | 4982123 | 167 | 29833.07 | 436724 | 918507 | 271.017 | 147.531 | 11.8148 |
| NO_VV_18_11 | 534 | ERS5890632 | 4971254 | 178 | 27928.39 | 450859 | 744905 | 220.527 | 148.024 | 10.3071 |
| NO_VV_18_01 | 534 | ERS5890728 | 4967651 | 320 | 15523.91 | 130767 | 1207502 | 312.315 | 129.323 | 32.0266 |
| NO_VV_18_02 | 534 | ERS5890733 | 4928925 | 227 | 21713.33 | 431213 | 1330526 | 377.171 | 141.738 | 21.9846 |
| NO_VV_18_03 | 534 | ERS5890723 | 4962429 | 203 | 24445.46 | 333215 | 1084928 | 307.814 | 141.859 | 22.1973 |
| NO_VV_18_05 | 534 | ERS5890725 | 4931669 | 165 | 29888.9 | 333502 | 1031903 | 300.464 | 145.587 | 16.4905 |
| NO_VV_18_04 | 535 | ERS5890800 | 4947916 | 169 | 29277.61 | 400596 | 380987 | 112.627 | 147.81 | 11.2296 |
| SE_VV_18_03 | 536 | ERS5890633 | 4913018 | 268 | 18332.16 | 160710 | 3452420 | 979.215 | 141.816 | 20.2831 |
| SE_VV_18_04 | 537 | ERS5890727 | 5025044 | 223 | 22533.83 | 226159 | 2287089 | 648.342 | 141.739 | 20.4438 |
| SE_VV_18_09 | 538 | ERS5890801 | 5005214 | 164 | 30519.6 | 305804 | 3538537 | 988.566 | 139.686 | 22.7628 |
| SE_VV_18_11 | 538 | ERS5890805 | 4989246 | 162 | 30797.81 | 456888 | 2592338 | 738.138 | 142.369 | 19.5335 |
| SE_VV_18_10 | 539 | ERS5890808 | 4886578 | 465 | 10508.77 | 147117 | 2435150 | 693.834 | 142.462 | 19.613 |

[1]***Note:*** *The first abbreviation represents the country (DK – Denmark, FI – Finland, NO – Norway, SE – Sweden) and the second abbreviation the Vibrio species. (VA – Vibrio alginolyticus, VC – non-toxigenic V. cholerae, VP – Vibrio parahaemolyticus, VV – Vibrio vulnificus). The first number represents the isolation year and the second number denotes the isolate number.*

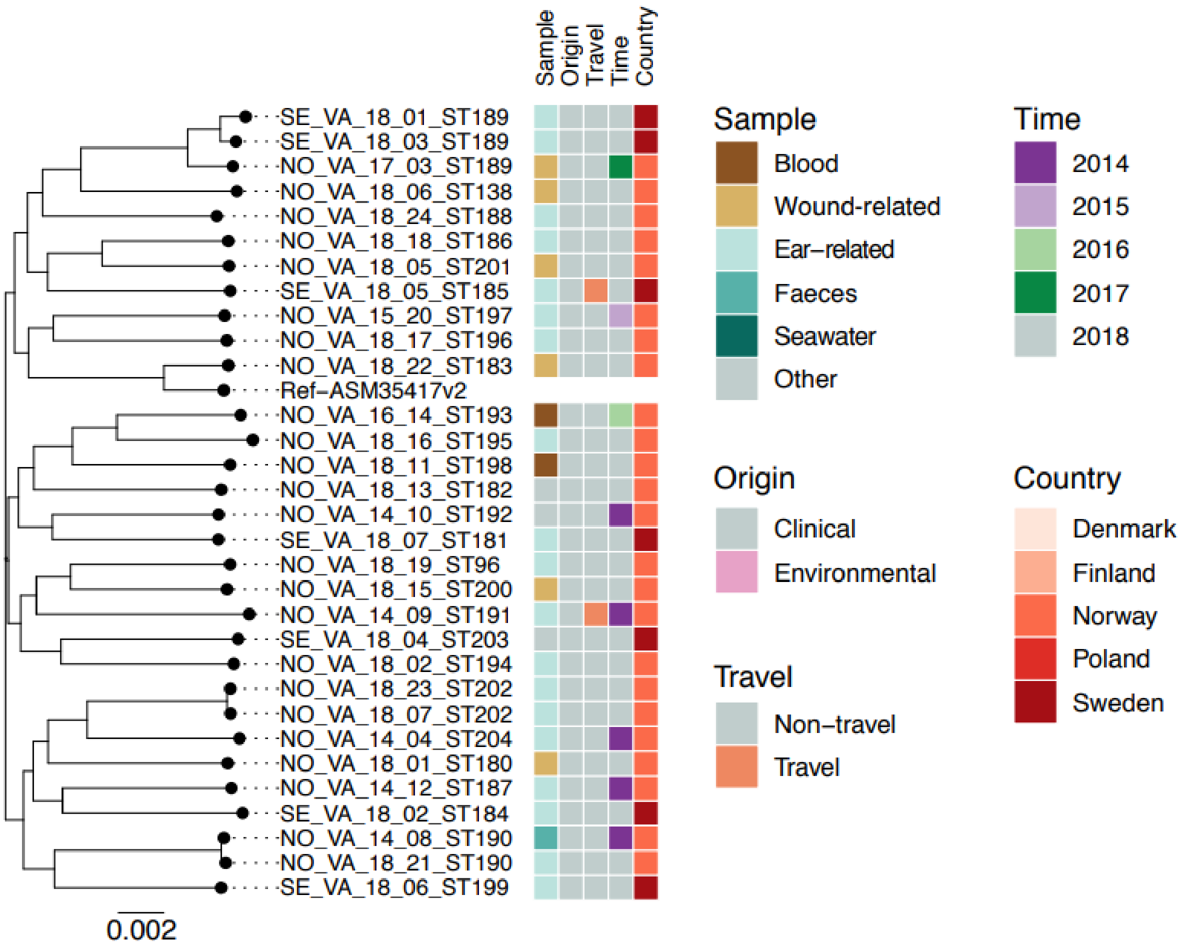


***Figure S1.*** *SNP based phylogeny of 31 V. alginolyticus phylogenetic genomes from study countries.* The *V. alginolyticus* ASM35417v2 sequence was used as reference. The scale bar indicates the number of substitutions per site.

Note: VA, *V. alginolyticus*, DK – Denmark, FI – Finland, NO – Norway, SE – Sweden, ST – sequence type. The first number represents the isolation year and the second number denotes the isolate number.

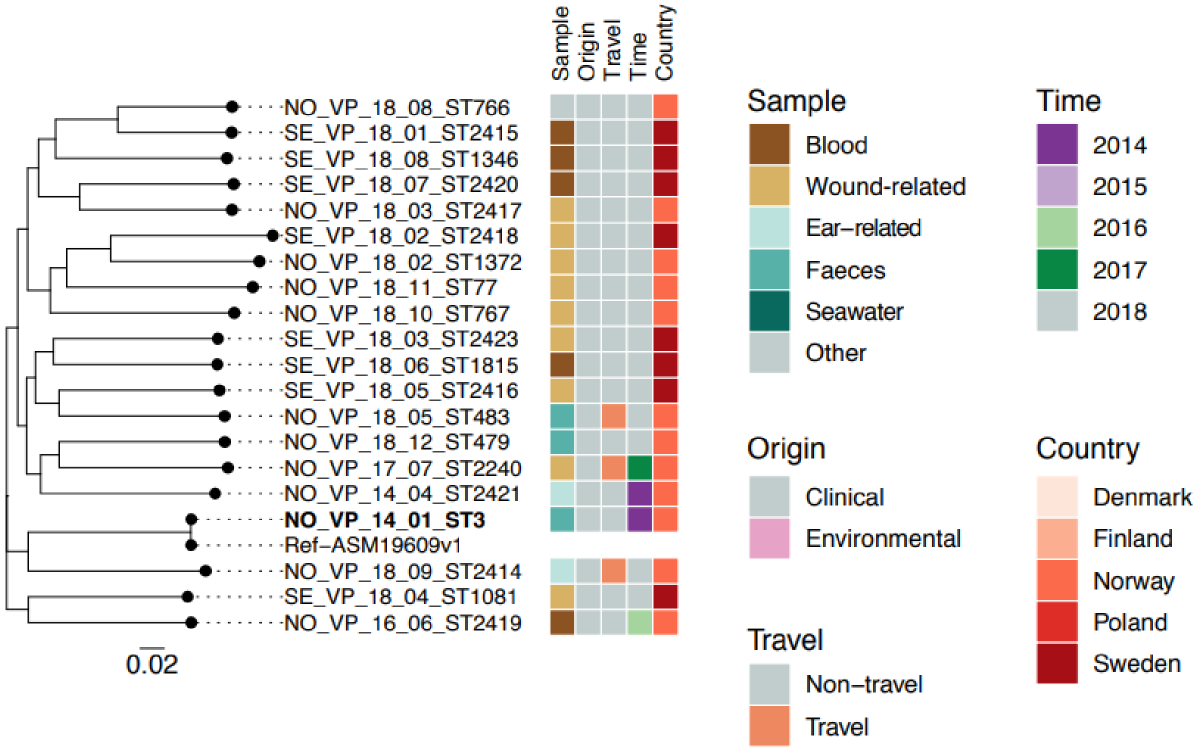


***Figure S2.** SNP based phylogeny of 20 V. parahaemolyticus genomes from study countries.* The *V. parahaemolyticus* ASM19609v1 sequence was used as reference. The scale bar indicates the number of substitutions per site.

Note: VP – *V. parahaemolyticus*, DK – Denmark, FI – Finland, NO – Norway, SE – Sweden, ST – sequence type. The first number represents the isolation year and the second number denotes the isolate number. Pandemic strain ST3 is bolded.
